# The value of brain age as a transdiagnostic biomarker of neurodegeneration

**DOI:** 10.64898/2026.07.21.26358569

**Authors:** Lonneke Bos, David R. van Nederpelt, James H. Cole, Bas Jasperse, Alle Meije Wink, Mario Tranfa, Eva Strijbis, Joep Killestein, Bernard M.J. Uitdehaag, Frederik Barkhof, Hugo Vrenken, Menno M. Schoonheim, Giuseppe Pontillo

## Abstract

Progressive structural brain changes are a hallmark of neurodegenerative conditions like Alzheimer’s disease (AD), frontotemporal dementia (FTD), multiple sclerosis (MS), and Parkinson’s disease (PD). The brain-predicted age difference (brain-PAD) has emerged as a promising biomarker to quantify these alterations, yet its unique clinical contribution relative to conventional measures of global brain atrophy such as the brain parenchymal fraction (BPF) remains underexplored.

In this transdiagnostic study across AD, FTD, MS, and PD, we systematically evaluated brain-PAD’s capacity to distinguish patients from controls, its cross-sectional and longitudinal associations with cognition, and its voxel-wise structural correlates. We benchmarked brain-PAD against BPF to determine its added explanatory value.

Brain-PAD successfully distinguished patients from controls, adding to BPF alone, in AD, FTD, and MS, but not PD. Across disorders, higher brain-PAD correlated with worse cognition, showing clear added value beyond BPF particularly in AD and MS. Baseline brain-PAD also independently predicted subsequent cognitive changes in AD, FTD, and MS, over and above BPF. Voxel-wise analyses revealed spatial features underlying brain-PAD including, beyond global tissue loss, specific regional atrophy matching each disease’s characteristic pattern.

Collectively, these findings demonstrate that brain-PAD is a clinically meaningful, transdiagnostic biomarker of neurodegeneration that complements conventional volumetric measures like the BPF.

## Introduction

Neurodegenerative diseases, including Alzheimer’s disease (AD) (1), frontotemporal dementia (FTD) (2), and Parkinson’s disease (PD) (3), are characterized by progressive structural changes in the brain. Similarly, while primarily classified as a neuroinflammatory disease, multiple sclerosis (MS) features a prominent and clinically relevant neurodegenerative component that makes it similar to primary neurodegenerative disorders (4). Across these conditions, quantifying brain alterations is essential for understanding disease mechanisms, tracking progression, and potentially evaluating treatment effects (5).

Brain volume measurements, derived from structural magnetic resonance imaging (MRI) scans, have long been used as markers of ageing and neurodegeneration (6–10). In particular, the brain parenchymal fraction (BPF), defined as the ratio of total brain volume over intracranial volume (ICV), is traditionally considered as a measure of lifetime brain atrophy, obtainable from a single cross-sectional MRI scan (11). As such, the BPF captures inter-subject variability associated with both non-pathological ageing and brain (neurodegenerative) diseases (12, 13).

In recent times, the brain-age paradigm has emerged as a promising approach for summarizing complex neuroimaging data into a single biomarker of ageing and neurodegeneration (14). Using machine learning, chronological age is modelled as a function of brain MRI scans in healthy people, and the resulting model of normal brain ageing is used for neuroimaging-based age prediction in unseen subjects. The extent to which a subject deviates from healthy brain ageing, expressed as the difference between predicted and chronological age (the brain-predicted age difference, brain-PAD), has been proposed as an age-adjusted global index of brain health, sensitive to a wide spectrum of neuropsychiatric conditions (15–18). Unlike the BPF, indexing how much brain tissue an individual currently has relative to maximal lifetime brain growth (proxied by ICV) (11), the “brain age” approach contextualizes individual brain appearance against a reference lifespan dataset, which makes it akin to normative modelling (19). In this framework, brain disorders are conceptualized as deviations from normal brain ageing (14). Also, the multivariate nature of brain age models, often operating directly on raw or minimally preprocessed structural MRI scans, makes them potentially sensitive to subtle, complex, and spatially distributed patterns of brain change that traditional univariate metrics might overlook (20).

Numerous studies have shown that patients with neurodegenerative conditions show a higher brain-PAD (i.e., their brains look older than expected based on their chronological age), which is associated with more severe clinical outcomes. In AD and FTD, higher brain-PAD is associated with worse cognitive performance (21). In MS, brain-PAD correlates with motor and cognitive disability (22–24). In PD, higher brain-PAD relates to greater motor and functional impairment (25). Beyond cross-sectional associations, longitudinal studies suggest that higher brain-PAD at baseline is associated with future clinical outcomes, including faster progression to dementia, worsening disability, and even increased mortality (17, 26–29). Together, these findings indicate that brain-PAD captures clinically relevant neurodegeneration spanning cognitive, motor, and functional domains, highlighting its potential as a generalizable biomarker across neurodegenerative disorders.

However, beyond the association with outcomes of interest, the path to clinical implementation requires demonstrating its added value over established measures and, ultimately, its clinical utility (30). In this regard, the unique contribution of brain-PAD relative to more conventional cross-sectional measures of lifetime brain atrophy, such as the BPF, remains underexplored. Furthermore, because brain age models collapse complex spatial features into a single metric, it remains unclear whether brain-PAD merely mirrors non-specific global atrophy or captures distinct neurodegenerative processes. A transdiagnostic evaluation of brain-PAD, in relation to BPF, across different etiologies could provide a more comprehensive account of its disease-specific sensitivity to clinical status and underlying patterns of brain structural loss.

In this study, we aimed to assess the value of brain-PAD, benchmarked against BPF, as a marker of neurodegeneration in AD, FTD, MS, and PD. First, we evaluated the capacity of brain-PAD to distinguish patients from healthy controls and provide additional explanatory power over BPF. Second, we examined the relationship between brain-PAD and disease-specific cognitive measures and determined whether brain-PAD accounts for variance in cognitive performance over and above that explained by BPF. Third, we assessed the association between brain-PAD and future cognitive worsening, and whether it explains longitudinal progression above and beyond BPF. Finally, we investigated the voxel-wise structural correlates of brain-PAD to assess whether the patterns of neurodegeneration associated with higher brain-PAD are disease-specific.

## Results

### Study population

We retrospectively collected and analysed 3 Tesla structural brain MRIs and clinical-demographic data from multiple cohorts. The final study population consisted of: 109 patients with Alzheimer’s disease (AD), 195 with mild cognitive impairment (MCI), and 185 cognitively normal (CN) participants from the Alzheimer’s Disease Neuroimaging Initiative (ADNI) cohort (31); 112 patients with frontotemporal dementia (FTD) (50 behavioural variant – BV-FTD, 31 progressive non-fluent aphasia – PNFA-FTD, 31 semantic variant – SV-FTD), and 129 CN participants from the Neuroimaging in Frontotemporal Dementia dataset (NIFD) cohort (32); 532 people with multiple sclerosis (pwMS) and 199 healthy controls (HCs) from the Amsterdam MS Cohort (MSCA) (33, 34); 373 patients with Parkinson’s disease (PD) and 121 HCs from the Parkinson’s Progression Markers Initiative (PPMI) cohort (35). When multiple sessions were available, we used the first MRI scan and modelled longitudinal clinical changes with all visits. Demographic, clinical, and MRI characteristics of the studied population are presented in Table 1.

**Table 1.**
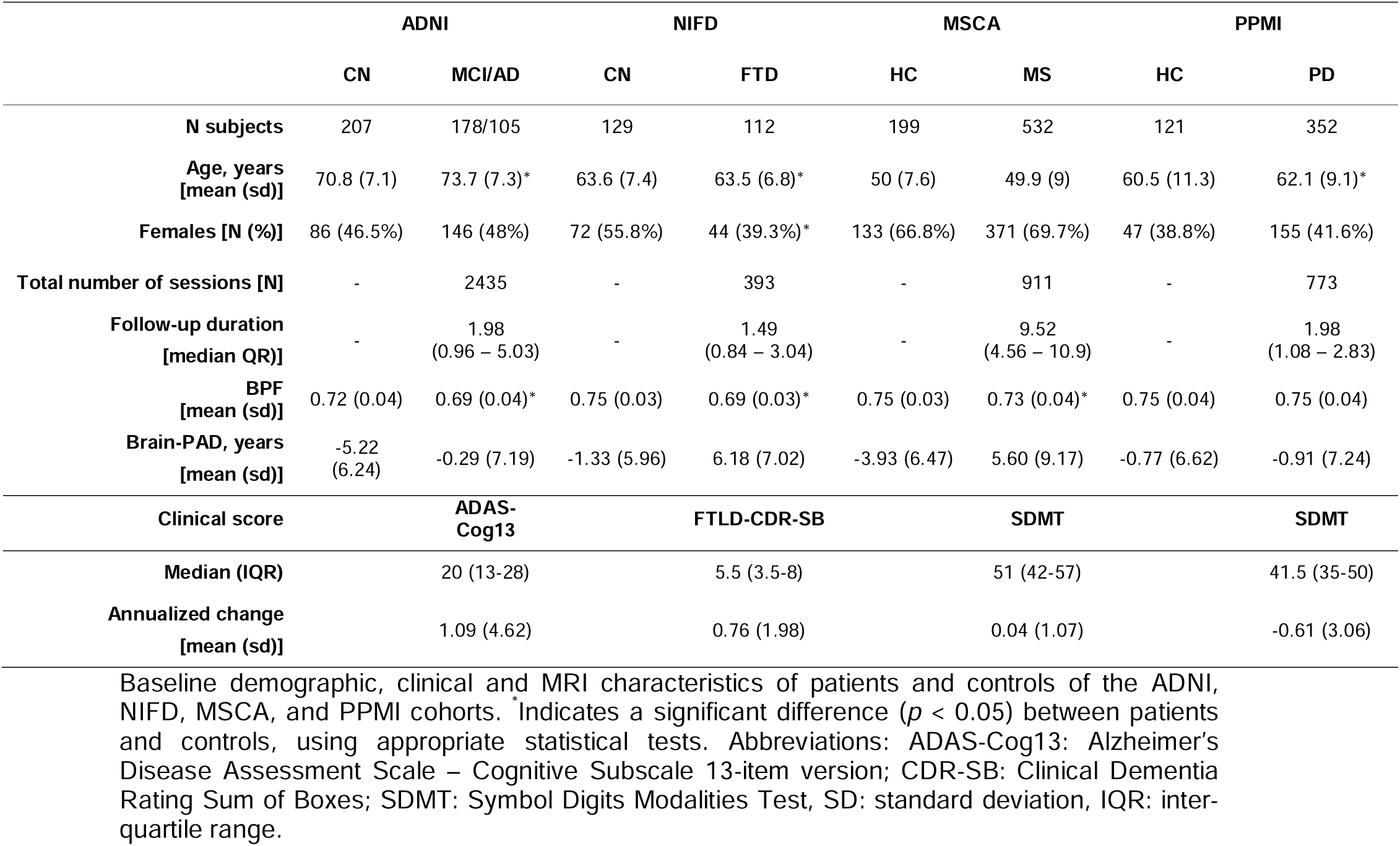
Demographic, clinical, and MRI characteristics of study cohorts.

We processed 3D T1-weighted (T1w) scans using FreeSurfer’s cross-sectional recon-all pipeline to obtain brain tissue volumes, from which we calculated BPF as the ratio of whole brain volume to estimated total intracranial volume (eTIV) (36). In addition, we obtained 3D T1w-based age predictions using the widely adopted *brainageR* model (15, 17, 37), and calculated brain-PAD as the difference between predicted and chronological age. Compared to controls, patients consistently showed lower BPF and higher brain-PAD values, except for patients with PD. Similarly, all patients except for those with PD showed older-looking brains than controls, with the greatest difference observed at the MS vs. HC comparison (9.52 years; 95% CI: 8.13, 10.90) (Table 1 and Figure 1).

**Figure 1.**
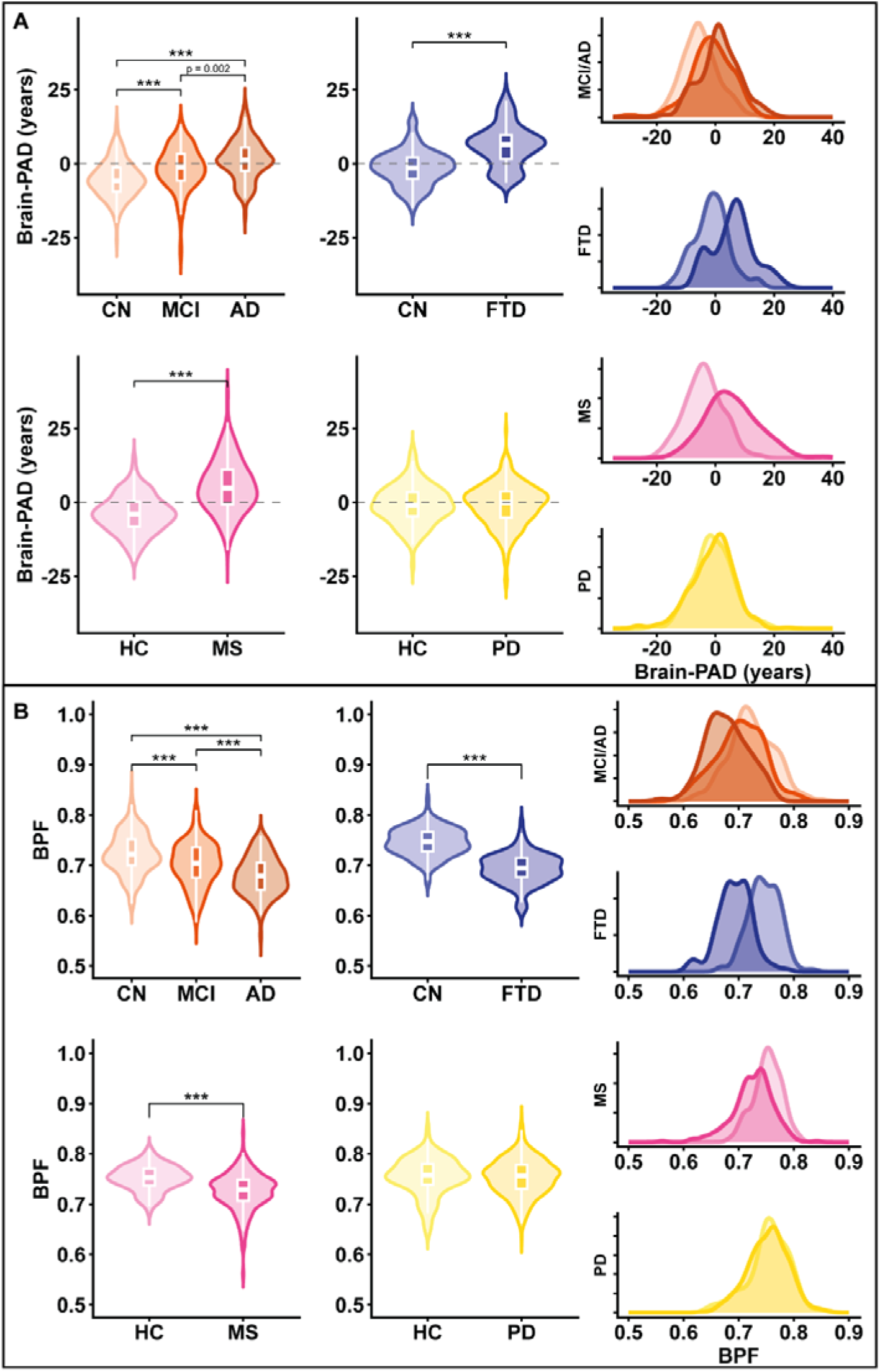
Distributions and between-group differences of brain-PAD and BPF across diseases. Violin plots showing the distribution of brain-PAD **(A)** and BPF **(B)** for each disease group (AD, FTD, MS and PD), where group differences were assessed using two-sided Wilcoxon signed rank-test. Boxes within the violin plots represent the median and IQR. Density plots display the overall distribution of brain-PAD and BPF, highlighting the range and skew of values across the cohorts. *** indicates *p*<0.001

### Brain-PAD discriminates patients from controls beyond BPF

We used logistic regression models to assess the value of brain-PAD and BPF for distinguishing between patients and controls across conditions, adjusting for age and sex. Brain-PAD was positively associated with the odds of belonging to the (more severe) patient group in the ADNI cohort (odds ratio, OR = 2.20; 95% CI: 1.81,2.68; *p* < 0.001), as well as for FTD (OR = 4.01; 95% CI: 2.75, 6.14; *p* < 0.001) and MS (OR = 4.38; 95% CI: 3.38, 5.77; *p* < 0.001), but not for PD (OR = 0.96; 95% CI: 0.81, 1.23, *p* = 0.96). Similarly, lower BPF was associated with increased chances of belonging to the (more severe) patient group in the ADNI cohort (OR = 2.55; 95% CI: 2.08, 3.16; *p* < 0.001) and for FTD (OR = 17.40; 95% CI: 9.07, 37.9; *p* < 0.001) and MS (OR = 2.77; 95% CI: 2.21, 3.54; *p* < 0.001), but not for PD (OR = 0.93; 95% CI: 0.73, 1.19; *p* = 0.56) (Figure 2A and Table 2).

**Figure 2.**
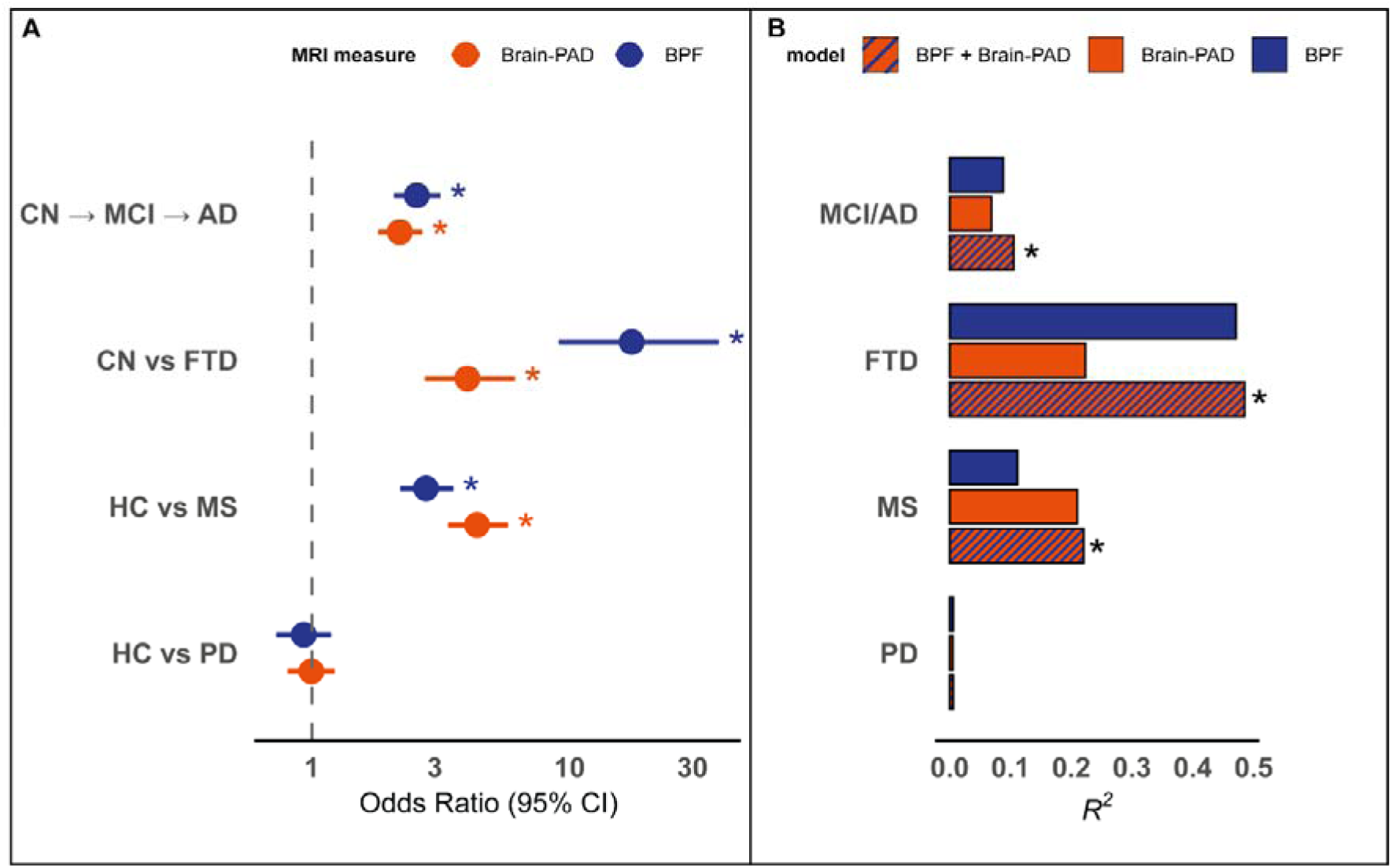
Discriminating patients from controls with brain-PAD and BPF across conditions. Panel **(A)** represents the odds ratios (ORs) and 95% CI from binary logistic regression models to discriminate patients from controls with either brain-PAD or BPF. ORs indicate the increased chances of belonging to the (more severe) patient group associated with one standard deviation change in the MRI measure. To ease interpretation, the sign of the BPF was flipped so that higher ORs consistently indicate a stronger positive association between MRI-based neurodegeneration (i.e., higher brain-PAD or lower BPF) and the odds of belonging to the (more severe) patient group. Panel **(B)** shows bar plots indicating the explained variance (*R*^2^) of models discriminating patients from controls using brain-PAD, BPF, or brain-PAD+BPF. All models were adjusted for age and sex. * indicates statistical significance.

**Table 2.**
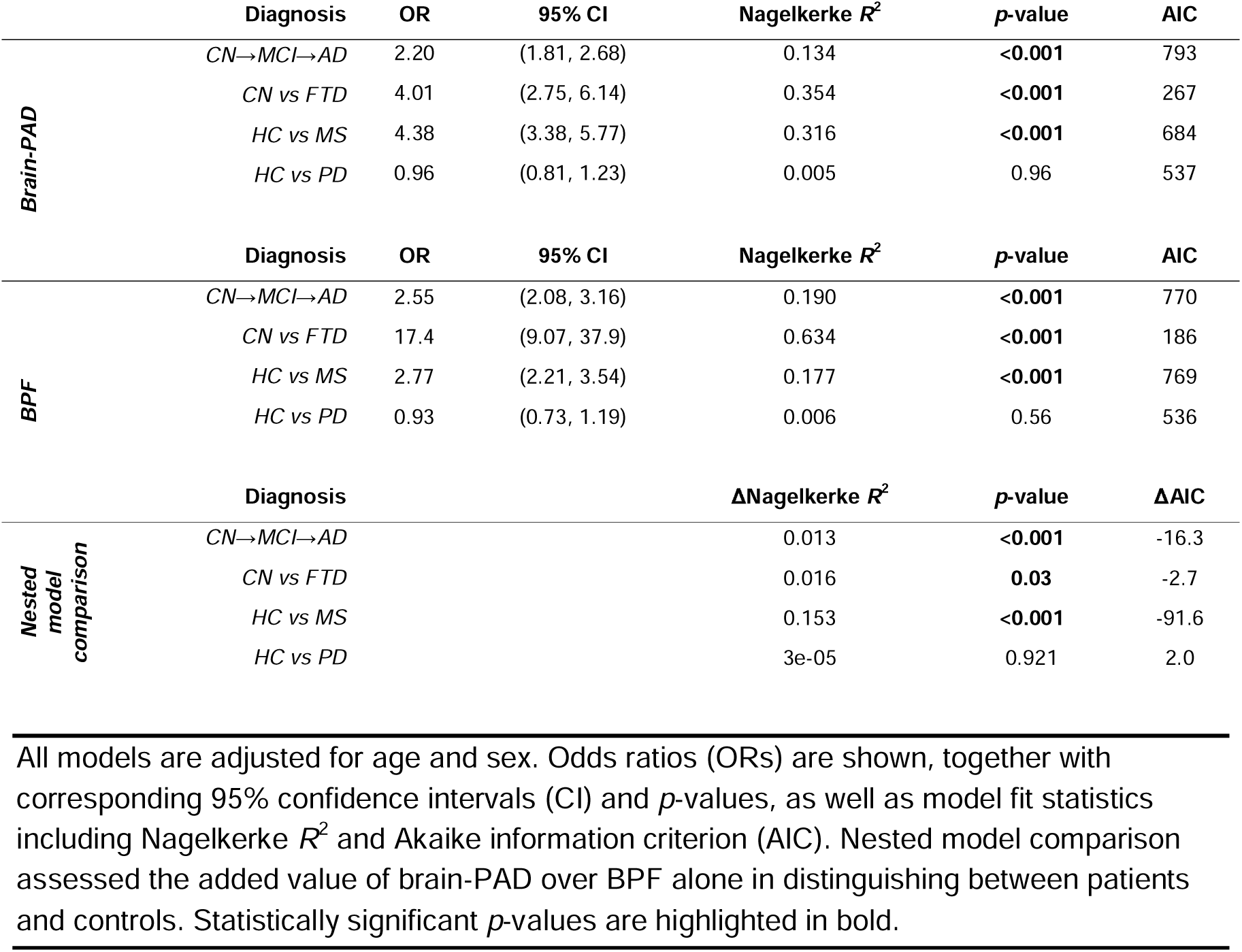
Logistic regression models discriminating (more severe) patient groups from controls using brain-PAD or/and BPF.

To determine whether brain-PAD explained diagnostic status beyond BPF, we assessed the added explanatory value of brain-PAD in a nested model comparison between a baseline logistic regression model including BPF as the main independent variable and an extended model additionally including brain-PAD. The extended models had significantly higher explanatory power, expressed with the Nagelkerke *R*^2^, in the ADNI cohort (Δ*R*^2^ = 0.013; *p* < 0.001), as well as for FTD (Δ*R*^2^ = 0.016; *p* = 0.03) and MS (Δ*R*^2^ = 0.153; *p* < 0.001), but not for PD (*p* = 0.92) (Figure 2B and Table 2).

### Brain-PAD explains cognitive performance beyond BPF

Using linear regression models including age and sex as covariates, we assessed the associations of brain-PAD and BPF with disease-specific cognitive measures: ADAS-Cog13 (Alzheimer’s Disease Assessment Scale - Cognitive Subscale, 13-item version) in MCI/AD, FTLD-CDR-SB (Frontotemporal Lobar Degeneration-modified Clinical Dementia Rating scale - Sum of Boxes) in FTD, and SDMT (Symbol Digit Modalities Test) in MS and PD. Higher brain-PAD was associated with worse cognitive performance in patients with MCI/AD (ADAS-Cog13: β = 0.333 ± 0.037; *p* < 0.001), FTD (FTLD-CDR-SB: β = 0.191 ± 0.081; *p* = 0.02), MS (SDMT: β = -0.397 ± 0.034; *p* < 0.001), and PD (SDMT: β = -0.122 ± 0.035; *p* < 0.001). Similarly, lower BPF was associated with worse cognitive performance in MCI/AD (ADAS-Cog13: β = -0.435 ± 0.039; *p* < 0.001), FTD (FTLD-CDR-SB: β = -0.501 ± 0.084; *p* < 0.001), and MS (SDMT: β = 0.427 ± 0.034; *p* < 0.001), but not in PD (SDMT: β = 0.027 ± 0.041; *p* = 0.52) (Table 3 and Figure 3A).

**Figure 3.**
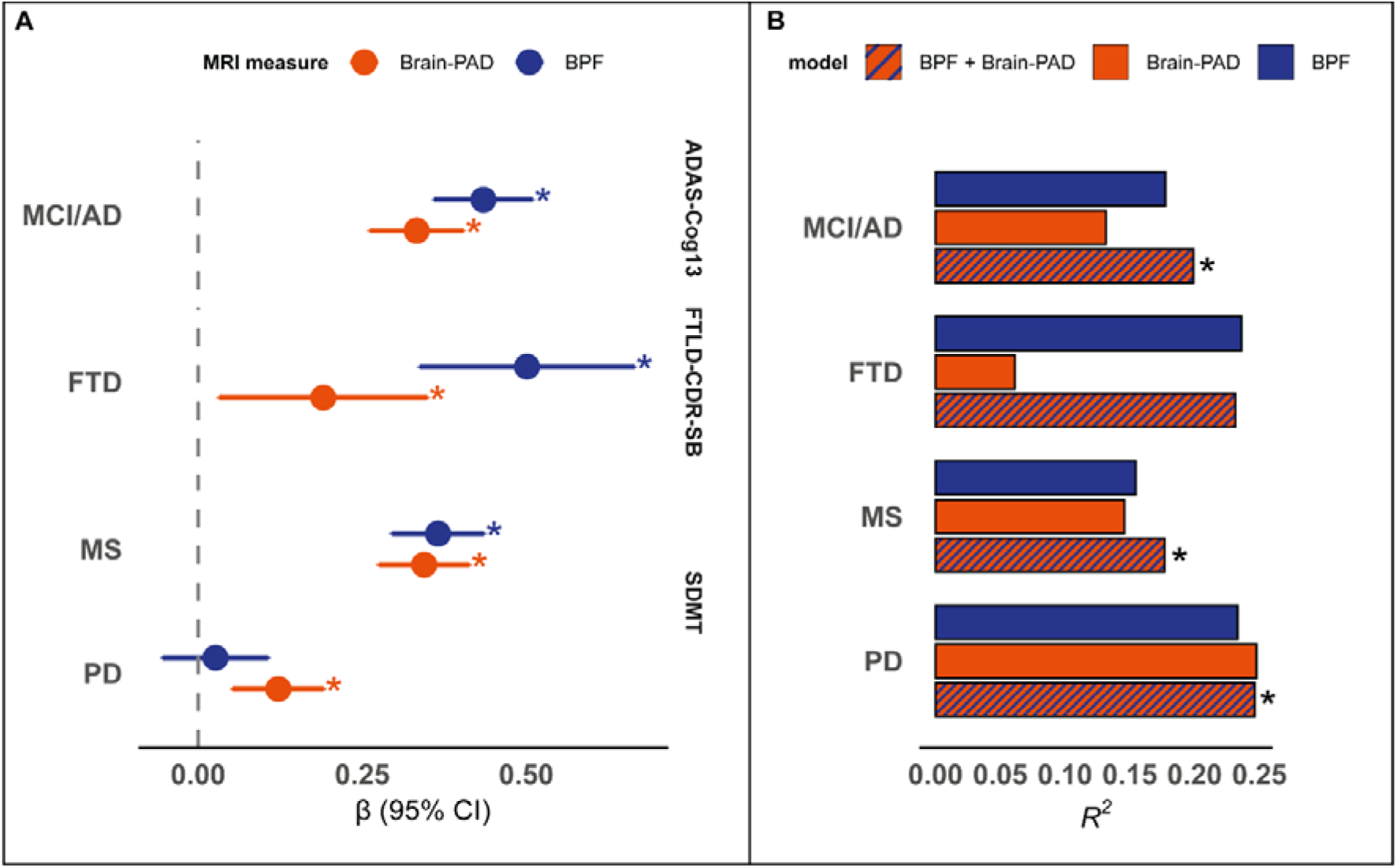
Associations with cognitive performance of brain-PAD and BPF across conditions. Panel **(A)** shows standardized betas (β) and 95% confidence intervals relative to the association between brain-PAD or BPF and cognitive performance measured by the ADAS-Cog13 in AD, FTLD-CDR-SB in FTD, and SDMT in MS and PD. β values indicate changes in cognitive performance (in standard deviations) per one standard deviation change in the MRI-derived variable. To ease interpretation, signs of BPF and SDMT were flipped so that higher βs consistently indicate a stronger positive association between MRI-based neurodegeneration (i.e., higher brain-PAD or lower BPF) and cognitive severity. Panel **(B)** shows bar plots indicating the explained variance (*R*^2^) of models explaining cognitive performance using brain-PAD, BPF, or brain-PAD+BPF. All models were adjusted for age and sex. * indicates statistical significance.

**Table 3.** Linear regression models explaining cognitive performance based on brain-PAD or/and BPF.

| | *Diagnosis* | *Cognitive test* | $\beta$ | SE | $R^2$ | *p-value* | AIC |
| --- | --- | --- | --- | --- | --- | --- | --- |
| *Brain-PAD* | *MCI/AD* | ADAS-Cog13 | 0.333 | 0.037 | 0.132 | **<0.001** | 1903 |
|  | *FTD* | FTLD-CDR-SB | 0.191 | 0.081 | 0.061 | **0.02** | 315 |
|  | *MS* | SDMT | -0.397 | 0.034 | 0.211 | **<0.001** | 1786 |
|  | *PD* | SDMT | -0.122 | 0.035 | 0.248 | **<0.001** | 1546 |
| | *Diagnosis* | *Cognitive test* | $\beta$ | SE | $R^2$ | *p-value* | AIC |
| *BPF* | *MCI/AD* | ADAS-Cog13 | -0.435 | 0.039 | 0.177 | **<0.001** | 1942 |
|  | *FTD* | FTLD-CDR-SB | -0.501 | 0.084 | 0.236 | **<0.001** | 343 |
|  | *MS* | SDMT | 0.427 | 0.034 | 0.227 | **<0.001** | 1800 |
|  | *PD* | SDMT | 0.027 | 0.041 | 0.233 | 0.52 | 1557 |
| | *Diagnosis* | *Cognitive test* | | | $\Delta R^2$ | *p-value* | $\Delta AIC$ |
| *Nested model comparison* | *MCI/AD* | ADAS-Cog13 |  |  | 0.022 | **<0.001** | -18.0 |
|  | *FTD* | FTLD-CDR-SB |  |  | -0.005 | 0.66 | 2.0 |
|  | *MS* | SDMT |  |  | 0.028 | **<0.001** | -24.0 |
|  | *PD* | SDMT |  |  | 0.237 | **<0.001** | -9.0 |

All models are adjusted for age and sex. Standardized betas ( $\beta$ ) are shown together with corresponding standard error (SE) and $p$ -values, as well as model fit statistics including adjusted $R^2$ and Akaike information criterion (AIC). Nested model comparison assesses the added value of brain-PAD over BPF alone in explaining cognitive performance. Statistically significant $p$ -values are highlighted in bold.

In a nested model comparison, brain-PAD provided additional explanatory value for cognitive performance beyond BPF in MCI/AD (extended vs. baseline model: Δ*R*² = 0.022; *p* < 0.001), MS (Δ*R*² = 0.028; *p* < 0.001) and PD (Δ*R*² = 0.237; *p* < 0.001), but not in FTD (Δ*R*² = −0.005; *p* = 0.66) (Table 3 and Figure 3B).

### Brain-PAD explains future cognitive decline beyond BPF

When modelling longitudinal cognitive changes at the group level, we observed significant cognitive worsening in patients with MCI/AD (β = 1.71; p < 0.001) and FTD (β = 1.17; p < 0.001), but not in MS (β = 0.03; p = 0.69) or PD (β = −0.39; p = 0.28) (Supplementary Figure 1).

As for the relationship between baseline MRI measures and individual-level annualized cognitive change over time, higher brain-PAD showed associations with faster subsequent cognitive decline in MCI/AD (ADAS-Cog13: β = 0.274 ± 0.090; p < 0.001), FTD (FTLD-CDR-SB: β = 0.328 ± 0.103; p < 0.01), and MS (SDMT: β = -0.157 ± 0.062; p < 0.05), but not in PD (SDMT: β = -0.003 ± 0.014; *p* = 0.96). In contrast, baseline BPF was only associated with longitudinal cognitive changes in MCI/AD (ADAS-Cog13: β = -0.290 ± 0.085; *p* < 0.001), while no significant associations were observed in FTD (FTLD-CDR-SB: β = -0.243 ± 0.142; *p* = 0.09), MS (SDMT: β = 0.053 ± 0.066; *p* = 0.42), or PD (SDMT: β = 0.018 ± 0.076; *p* = 0.82) (Table 4 and Figure 4A).

**Figure 4.**
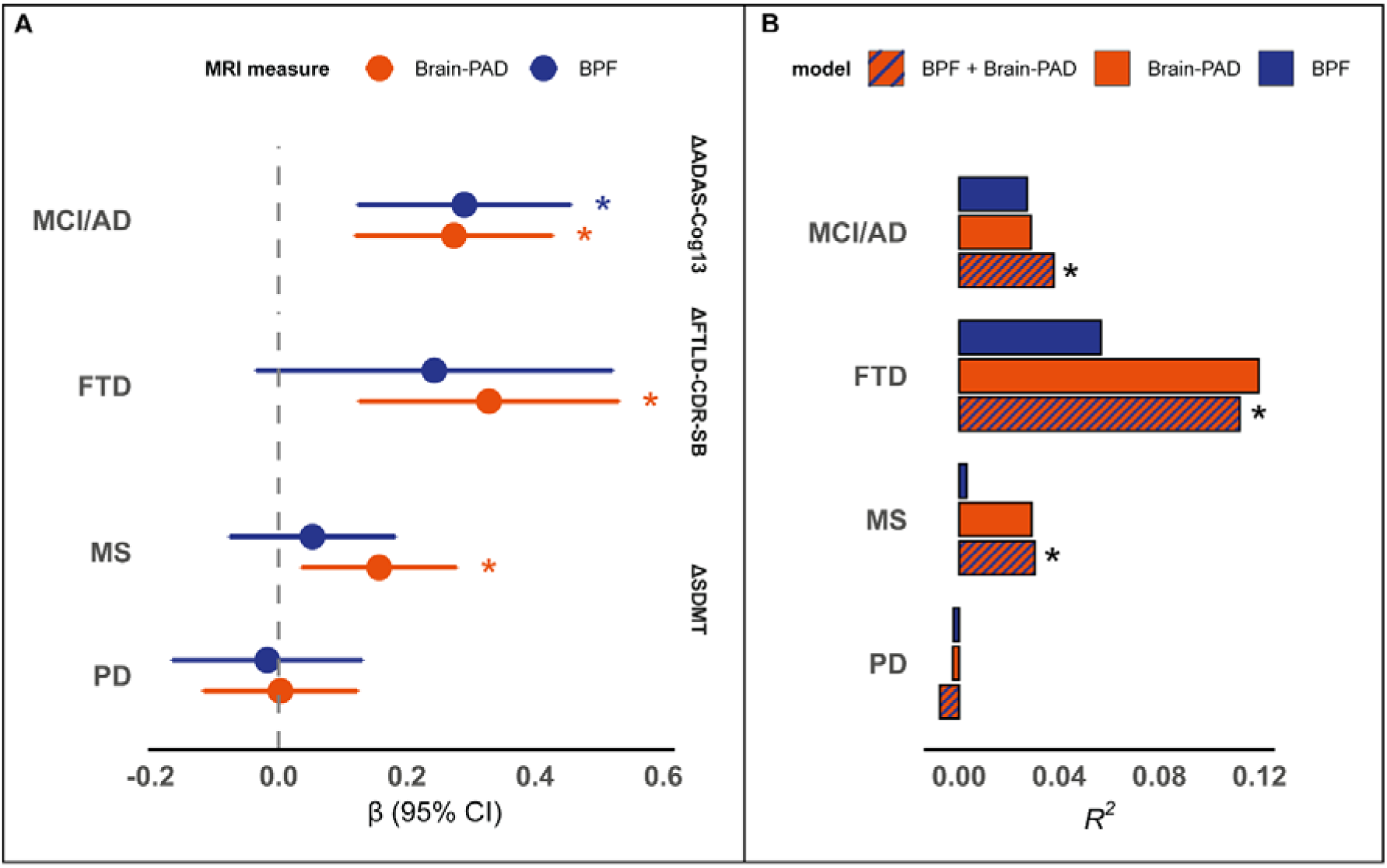
Associations with longitudinal cognitive change of brain-PAD and BPF across conditions. Panel **(A)** shows standardized betas (β) and 95% CI relative to the association between brain-PAD or BPF and annualized change in cognitive performance, measured by the ADAS-Cog13 in AD, FTLD-CDR-SB in FTD, and SDMT in MS and PD. β values indicate changes in cognitive progression (in standard deviations) per one standard deviation change in the MRI-derived variable. To ease interpretation, signs of BPF and SDMT were flipped so that higher βs consistently indicate a stronger positive association between MRI-based neurodegeneration (i.e., higher brain-PAD or lower BPF) and cognitive progression. Panel **(B)** shows bar plots indicating the explained variance (*R*^2^) of models explaining cognitive progression using brain-PAD, BPF, or brain-PAD+BPF. All models were adjusted for age and sex. * indicates statistical significance.

**Table 4.** Linear regression models explaining future cognitive changes based on brain-PAD or/and BPF.

| | Diagnosis | Clinical outcome | $\beta$ | SE | $R^2$ | $p$-value | AIC |
| --- | --- | --- | --- | --- | --- | --- | --- |
| Brain-PAD | MCI/AD | $\Delta ADAS-Cog13/y$ | 0.274 | 0.078 | 0.029 | **<0.001** | 1110 |
| | FTD | $\Delta CDR-SB/y$ | 0.328 | 0.103 | 0.12 | **0.002** | 310 |
| | MS | $\Delta SDMT/y$ | -0.157 | 0.062 | 0.029 | **0.01** | 592 |
| | PD | $\Delta SDMT/y$ | -0.003 | 0.014 | -0.002 | 0.96 | 485 |
| | Diagnosis | Clinical outcome | $\beta$ | SE | $R^2$ | $p$-value | AIC |
| BPF | MCI/AD | $\Delta ADAS-Cog13/y$ | -0.290 | 0.085 | 0.027 | **<0.001** | 1111 |
| | FTD | $\Delta CDR-SB/y$ | -0.243 | 0.142 | 0.057 | 0.09 | 317 |
| | MS | $\Delta SDMT/y$ | 0.053 | 0.066 | 0.003 | 0.42 | 597 |
| | PD | $\Delta SDMT/y$ | 0.018 | 0.076 | -0.002 | 0.82 | 485 |
| | Diagnosis | Clinical outcome | | | $\Delta R^2$ | $p$-value | $\Delta AIC$ |
| Nested model comparison | MCI/AD | $\Delta ADAS-Cog13/y$ | | | 0.038 | **0.04** | -2.5 |
| | FTD | $\Delta CDR-SB/y$ | | | 0.112 | **0.009** | -5.2 |
| | MS | $\Delta SDMT/y$ | | | 0.030 | **0.008** | -5.1 |
| | PD | $\Delta SDMT/y$ | | | 0.005 | 0.90 | 2.0 |

All models are adjusted for age and sex. Standardized betas ( $\beta$ ) are shown, together with corresponding standard error (SE) and $p$ -values, as well as model fit statistics including adjusted $R^2$ and Akaike Information Criterion (AIC). Nested model comparison assesses the added value of brain-PAD over BPF alone in explaining annualized cognitive change. Statistically significant $p$ -values are highlighted in bold.

In a nested model comparison, brain-PAD provided additional explanatory value beyond BPF for subsequent cognitive changes in MCI/AD (Δ*R*² = 0.038; *p* = 0.04), FTD (Δ*R*² = 0.112, *p* = 0.009), and MS (Δ*R*² = 0.030; *p* < 0.008), but not in PD (Δ*R*² = 0.005; *p* = 0.90) (Table 4 and Figure 4B).

### Sensitivity analyses using alternative brain age models

To assess the robustness of our findings to the choice of the brain age framework, we conducted sensitivity analyses using alternative, publicly available state-of-the-art deep learning models: *DeepBrainNet* (38), the *MIDI-model* (20), and *pyment* (39).

The capacity of brain-PAD to distinguish patients with MCI/AD, FTD, or MS from controls, alone and in addition to BPF, was consistent with *DeepBrainNet* and, partially, with the *MIDI-model*, but not with *pyment*, which was only sensitive to MS and PD (Supplementary Figure 2).

For cross-sectional cognitive measures, the associations observed with *brainageR* were substantially replicated with *DeepBrainNet*, though with a lower effect size in MS, and with the *MIDI-model*. Mirroring the diagnostic classification analysis, *pyment* proved to be the least sensitive, significantly explaining cognition, alone and in addition to BPF, only within the MS and PD groups (Supplementary Figure 2).

As for the relationship with future cognitive change, the primary results were only partially replicated with *DeepBrainNet*, demonstrating that higher baseline brain-PAD and was associated with faster clinical decline specifically in the MCI/AD patient group. Conversely, no significant associations between brain-PAD and future cognitive progression across conditions were observed with the *MIDI-model* or *pyment* (Supplementary Figure 2).

### Neuroanatomical correlates of brain-PAD

To investigate whether brain-PAD was associated with disease-specific neuroanatomical patterns across conditions, we conducted voxel-based morphometry (VBM) analyses in each diagnostic group (Figure 2 and Supplementary Figures 3 and 4). In the HC group no clear regional specificity emerged, with brain-PAD showing a widespread negative relationship with brain tissue volumes. In MCI/AD, the strongest negative relationship between brain-PAD and brain tissue volumes was observed in fronto-temporal regions, and the weakest in the occipital lobe. Despite the low sample size, distinct variant-specific patterns were observed in FTD, with higher brain-PAD mostly associated with frontal atrophy in patients with BV-FTD, fronto-temporal atrophy in patients with PNFA-FTD, and temporal pole atrophy in patients with SV-FTD. In MS, there was a strong negative relationship between brain-PAD values and brain tissue volumes in the deep white matter (WM) and subcortical gray matter (GM), as well as in the Rolandic cortex (Figure 3). In PD, there was a slight focus on subcortical areas, although no strong regional specificity was observed.

When repeating the voxel-wise analyses after including BPF as an additional covariate, we observed a general reduction of the magnitude of associations between brain-PAD and local tissue volumes, but their spatial distribution remained mostly unaltered, suggesting that brain-PAD is sensitive to disease-specific neuroanatomical patterns beyond global brain atrophy (Supplementary Figure 5).

**Figure 3.**
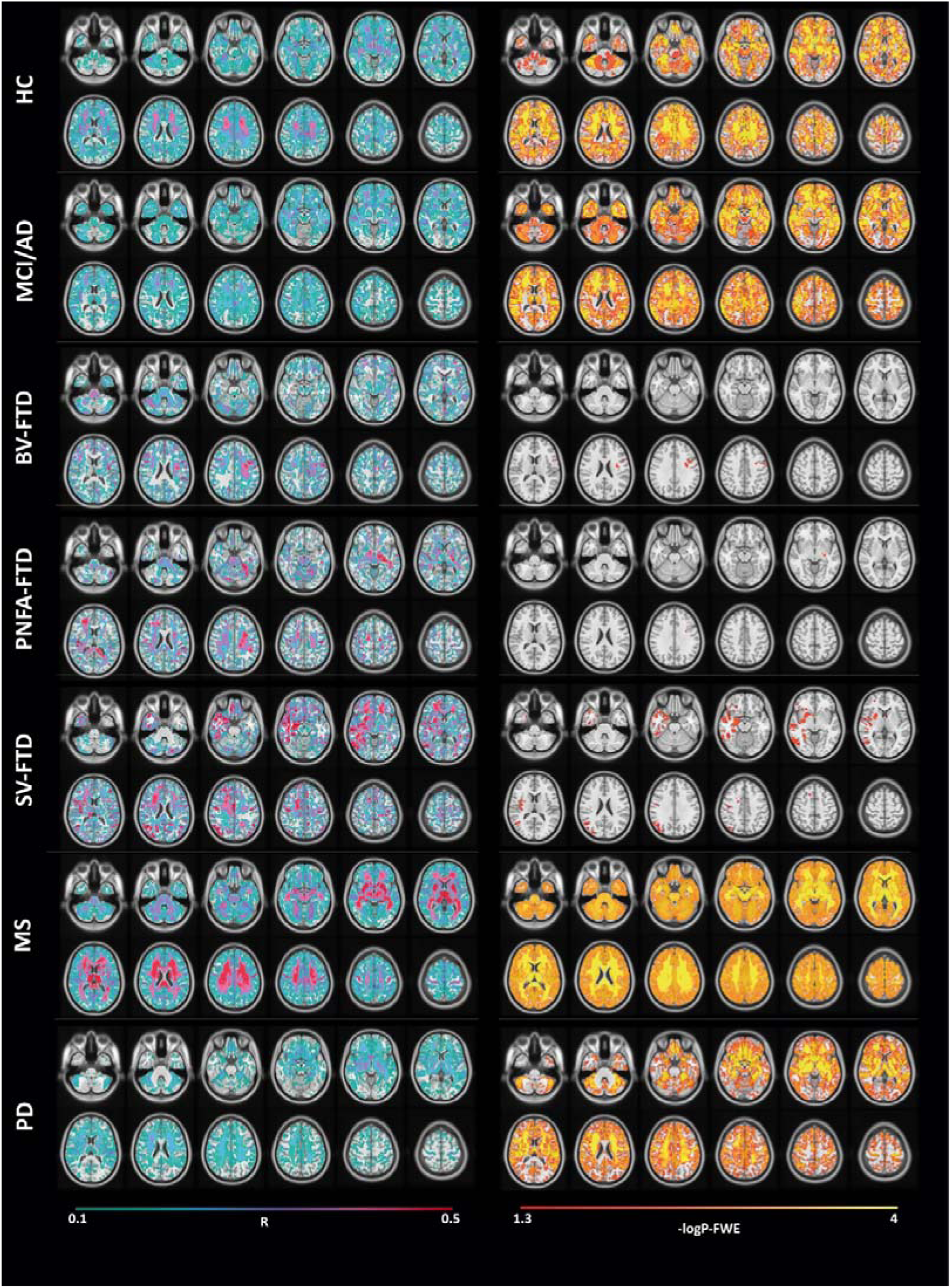
Voxel-wise associations between brain-PAD and brain tissue volumes. Effect size (R, *sea green* to *red*) and thresholded statistical (−logP-FWE, *red* to *yellow*) maps of the negative relationship between brain-PAD and voxel-wise tissue volume are shown across diagnostic groups, superimposed on axial sections of a 3D T1-weighted template in standard space. All analyses were adjusted for age, sex, and total intracranial volume.

## Discussion

In this study, we systematically evaluated brain-PAD, in relation to BPF, as a marker of neurodegeneration across MCI/AD, FTD, MS, and PD. We found that brain-PAD was consistently associated with disease status, cognition and future cognitive decline in MCI/AD, FTD, and MS, providing additional explanatory value beyond BPF. Voxel-wise analyses revealed distinct, disease-specific neuroanatomical correlates of brain-PAD, partially independent of BPF. Overall, our findings highlight the potential of brain-PAD as a sensitive transdiagnostic marker of neurodegeneration, complementing conventional measures of lifetime brain atrophy.

As expected, both brain-PAD and BPF were sensitive to MCI/AD, FTD, and MS, all diseases characterized by substantial brain atrophy since the early phases. Across these conditions, brain-PAD added to BPF for discriminating patients from controls, suggesting that the two metrics capture distinct, yet partially overlapping, aspects of neurodegeneration. The greatest added value of brain-PAD over BPF was observed in MS, which might be explained by the different spatial distribution of pathology across these conditions. While FTD and AD primarily cause prominent cortical thinning, MS is characterized by early and pronounced subcortical damage. Due to its geometry and histological organization, cell damage/loss within the cerebral cortex translates directly into large volumetric drops (40), which are captured by a global macrostructural metric like BPF. Conversely, pathological changes in subcortical compartments, which are compact and highly connected, tend to alter complex structural networks and spatial covariance patterns before manifesting as a crude loss of global parenchymal volume. Multivariate brain age models are inherently optimized to detect these subtle, distributed spatial patterns, making them potentially more sensitive to subcortical damage.

In contrast, neither metric was sensitive to PD. This is consistent with the neurobiological profile of early-stage PD, featuring subtle, regionally confined pathology rather than widespread neurodegeneration. Indeed, the PPMI cohort restricts enrollment to patients within 2 years of diagnosis, when macrostructural brain changes are minimal. When investigating patients in more advanced phases of the disease, other studies reported a slight brain-PAD increase (ranging from 0.7 to 4.4 years) in PD (41–44).

Within each disease group, a higher brain-PAD was associated with poorer cognitive performance, in line with previous literature (21, 22, 25). Although BPF tended to account for a greater proportion of the variance in cognitive scores than brain-PAD, combining the two metrics provided additional explanatory value over BPF alone in both the MCI/AD and MS groups. As for their prognostic value, higher baseline brain-PAD was associated with future cognitive worsening in all conditions except for PD, outperforming and adding to BPF. While BPF reflects cumulative lifetime atrophy and therefore captures both physiological and disease-related inter-individual variability, the “normative” aspect of brain-PAD isolates deviations from healthy ageing, which might make it more sensitive to the disease momentum that drives future cognitive worsening. Ultimately, combining the two metrics provided additional explanatory value, confirming their complementary nature.

Importantly, while the complementarity of these two accessible structural markers is appealing for clinical translation, the added value of brain-PAD over BPF tended to be statistically significant but often marginal across our experiments. This suggests that a major portion of clinically relevant neurodegeneration is fundamentally captured by global brain atrophy. Consequently, a crucial direction for future research is to move beyond simple statistical associations and establish true clinical utility. In practice, this requires concrete evidence that implementing metrics like brain-PAD or BPF, alone or in combination, leads to a net positive impact on patient care, e.g., by enabling earlier and more accurate patient stratification and improving clinical decision-making.

We also investigated the impact of the brain age model on our analyses. While the main results were substantially replicated with *DeepBrainNet*, the *MIDI-model* and, particularly, *pyment* were less sensitive to disease status and cognitive variability, both cross-sectionally and longitudinally. Interestingly, *pyment* is also the model with the highest reported accuracy for chronological age prediction. This discrepancy likely reflect the previously described accuracy/utility trade-off for brain age models (45, 46). Specifically, more complex, accuracy-optimized brain age models may prioritize low-variance ageing features and therefore be less sensitive to (disease-related) inter-individual variance than “loose-fitting” models. Ultimately, these findings highlight that brain age estimates, and their clinical utility, are highly model-dependent (47, 48). This variability warrants caution when interpreting results and comparing findings across studies and underscores the need for standardization and further technical and clinical validation.

Voxel-wise analyses revealed that the neuroanatomical correlates of brain-PAD tended to recapitulate known disease-specific patterns of neurodegeneration. While no clear regional specificity emerged in controls, higher brain-PAD in MCI/AD was most strongly associated with lower frontal and temporal tissue volumes, with relative sparing of the occipital lobe. Within the FTD cohort, brain-PAD captured the signature neurodegeneration pattern for each variant. In patients with BV-FTD, the strongest negative association between brain-PAD and tissue volume occurred in frontal regions (49). In PNFA-FTD, higher brain-PAD was most strongly linked to fronto-temporal atrophy, with a slight left-sided predominance (50). In SV-FTD, we observed a strong association with temporal pole atrophy that was paradoxically more evident on the right side (51). This rightward skew was likely driven by a documented ceiling effect (52), wherein advanced neurodegeneration of the left temporal pole leaves insufficient structural variance across patients for VBM to detect. In MS, the strongest negative association between brain-PAD and brain tissue volume was observed in the deep WM, subcortical GM, and sensorimotor cortex. Finally, in patients with PD, higher brain-PD was most strongly associated with lower subcortical tissue volumes. While adjusting for BPF expectedly attenuated the magnitude of the statistical associations, their spatial topography remained substantially unaltered. Ultimately, these findings suggest that brain age captures region-specific structural change, providing information beyond global atrophy.

Our study has several limitations. First, the MS dataset is derived from a single center, which may limit generalizability due to site-specific acquisition and population characteristics. Multi-center validation studies are needed to confirm robustness across different scanners and clinical settings. Additionally, BPF was derived using different FreeSurfer versions, which may bias segmentation outputs, although the used version was consistent within cohorts (53). Finally, the used datasets are research cohorts rather than real-world data, which may limit generalizability to routine clinical populations. Future work should therefore focus on external validation in real-world settings to assess clinical feasibility and predictive utility.

In conclusion, brain-PAD captures clinically relevant and (partially) disease-specific patterns of neurodegeneration beyond global brain atrophy as measured with the BPF. While prospective validation is required to establish its direct clinical impact, our results support the value of brain-PAD as a robust transdiagnostic marker of neurodegeneration that effectively complements conventional volumetric measures.

## Methods

### Study cohorts

This study included data from four cohorts: 1) ADNI (31), 2) NIFD (32), 3) MSCA, containing Project Y (same-age cohort) and PrograMS (10-year follow-up longitudinal cohort) (33, 34), and 4) PPMI (35). We included all subjects labelled as AD, MCI, FTD, PD, and MS according to each study’s protocol, as well as HCs or CN subjects, with a high resolution (voxel size ≤ 1mm isotropic) 3D T1-weighted scan at 3T. When multiple sessions were available, we used the first MRI scan, while longitudinal clinical changes were modelled with all visits. In the case of ADNI, to ensure that clinical diagnoses were supported by underlying Alzheimer’s pathology and to mitigate the inclusion of spurious cases, we only included patients with MCI and dementia demonstrating AD biomarker positivity, defined as a CSF p-tau_181_/Aβ_42_ ratio > 0.021, as recommended by the ADNI Biomarker Core Steering Committee (54). Similarly, we only included CN subjects who were AD biomarker negative (CSF p-tau_181_/Aβ_42_ ratio ≤ 0.021). For the NIFD cohort, patients with behavioural variant (BV-FTD), progressive non-fluent aphasia (PNFA-FTD), and semantic variant (SV-FTD) were considered jointly as a single FTD group, except for voxel-wise analyses.

### Cognitive measures

We focused exclusively on cognitive measures as these are more directly related to brain neurodegeneration, whereas motor and functional assessments can be influenced by other factors (e.g., spinal cord damage in MS). Cognitive measures included the ADAS-Cog13 for AD patients (55), the FTLD-CDR-SB for FTD (56), and the SDMT for MS and PD (57).

### MRI processing

For ADNI, we used UCSF cross-sectional FreeSurfer data, processed with version 5.1 (58). For NIFD and PPMI, image processing was performed at our site using FreeSurfer version 6.0.1’s recon-all cross-sectional pipeline (36). For the MSCA data, we used a previously described pipeline (59) including MS lesion segmentation with nicMSlesions (60) and lesion filling with the lesion segmentation toolbox (LST) (61), followed by FreeSurfer version 7.3.2’s recon-all cross-sectional pipeline on lesion-filled images.

For ADNI data, we only retained scans which passed the visual QC according to UCSF FreeSurfer protocol. For locally processed data, we used the Euler number, an index of cortical surface reconstruction quality generated automatically by FreeSurfer. In particular, we discarded scans below an Euler number cutoff threshold of -120 (62).

We calculated the whole brain volume as the sum of the following structures: total cerebral GM, total cerebral WM, subcortical GM, brainstem, cerebellar GM, and cerebellar WM. BPF was obtained by dividing the whole brain volume by the estimated total intracranial volume (eTIV).

### Brain age prediction

We used 3D T1-weighted scans to predict age and calculated brain-PAD as the difference between brain-predicted and chronological age. Given its wide adoption in the field, we used *brainageR* as the primary brain age model. *brainageR* was trained on data from 3,377 healthy individuals (mean age ± SD: 40.6 ± 21.4 years; range: 18–92 years) across seven publicly available datasets and tested on an independent cohort of 857 healthy individuals (mean age ± SD: 40.1 ± 21.8 years; range: 18–90 years) (10, 21, 22). The reported mean absolute error (MAE) for this model was 4.9 years. Within *brainageR*’s framework, T1w scans are pre-processed with SPM12 (23) for segmentation and normalization to a template, then converted into feature vectors representing grey matter, white matter, and CSF, which are then reduced via principal component analysis (PCA). Principal components are used as input to a Gaussian Process Regression (GPR) model to predict age.

To assess the robustness of our findings to the choice of the brain age prediction model, we also replicated the analyses using alternative, publicly available, state-of-the-art deep learning models, namely *DeepBrainNet* (*38*), *MIDI-model* (20), and *pyment* (63). For details on these models’ architecture, training protocol, and performance, we refer the reader to the original publications.

### Statistical analyses

Statistical analyses were performed using R Statistical Software (version 4.3.2; R Foundation for Statistical Computing, Vienna, Austria). Assumptions for parametric tests were checked as appropriate, and a two-sided *p*-value < 0.05 was considered statistically significant. All statistical models were adjusted for age and sex.

#### Descriptives and between-group differences

Within each cohort, age and sex differences between patients and controls were tested using a linear model and the Chi-squared test, respectively. Differences between patients and controls in terms of brain-PAD and BPF were visualised using violin plots and density plots and tested with Wilcoxon signed-rank test.

#### Discriminating patients from controls

To assess the value of each biomarker in distinguishing patients from controls (CN vs FTD, HC vs MS and HC vs PD), we used binary logistic regression models. For the ADNI cohort, we used ordinal logistic regression to model disease progression from CN to MCI to AD, after preliminarily verifying the proportional odds assumption.

To determine whether brain-PAD explained diagnostic status beyond BPF, we evaluated the added explanatory value of brain-PAD in a nested model comparison between a baseline logistic regression with BPF as the main independent variable (eq. 1) and an extended model additionally including brain-PAD (eq. 2). Model comparisons were performed using a Chi-square test and, if significant, changes in explained variance (Δ*R*²) were calculated to quantify the added contribution of brain-PAD.

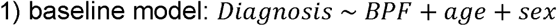

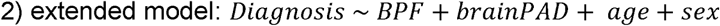

#### Explaining cognitive performance

We assessed the relationship between each imaging biomarker and disease-specific cognitive measures across cohorts. Specifically, we evaluated the relation between both brain-PAD and BPF with ADAS-Cog13 in AD, FTLD-CDR-SB in FTD, and SDMT scores in MS and PD, using linear regression models.

Subsequently, to determine whether brain-PAD explained clinical severity beyond BPF, we evaluated the added explanatory value of brain-PAD in a nested model comparison between a baseline linear regression including BPF as the main independent variable (eq. 3) and an extended model that additionally included brain-PAD (eq. 4). Model comparisons were performed using an F-test, and changes in explained variance (Δ*R*²) were calculated to quantify the added contribution of brain-PAD.

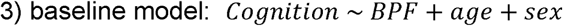

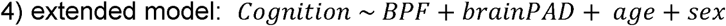

##### Explaining cognitive progression

For longitudinal analyses, we first modelled longitudinal cognitive changes at the group level with linear mixed-effects models with timepoints nested within patients and adjusting for age at baseline and sex. Then, we calculated participant-level annualized clinical changes by fitting individual linear regression models of cognitive measures against follow-up times (in years). Individual slopes were then used as the dependent variables in multiple linear regression analyses to examine associations with baseline brain-PAD or BPF.

To assess whether brain-PAD explained annualized cognitive change beyond BPF, we evaluated its added value in a nested model comparison between a baseline including BPF as the main independent variable (eq. 5) and an extended model additionally incorporating brain-PAD (eq. 6). Model comparisons were conducted using an F-test, and the added explanatory value of brain-PAD was quantified in terms of Δ*R*².

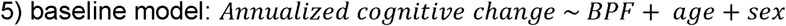

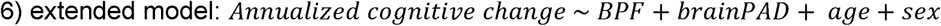

#### Voxel-Based Morphometry

To investigate the neuroanatomical correlates of brain-PAD across disorders, we processed 3D-T1w scans using DeepMRIPrep (64), a fully automated pipeline optimized for robust tissue segmentation and normalization, to generate normalized (i.e., to a common space), modulated, and smoothed (3mm FWHM Gaussian kernel) GM and WM maps. Afterwards, voxel-wise associations between pre-processed tissue maps and brain-PAD were assessed using a nonparametric approach based on 5000 permutations applied to the general linear model (65) via the Threshold Free Cluster Enhancement toolbox (66), while adjusting for age, sex, and TIV. Statistical analyses were constrained using GM and WM masks obtained by thresholding mean maps at 0.4 and 0.2 probability, respectively, to minimize overlap between tissue classes. To assess the unique relationships between local tissue volumes and brain-PAD, independent of global brain atrophy, the same analyses were repeated after adding BPF as an additional covariate. These analyses were conducted separately for each diagnostic group (MCI/AD, MS, PD, FTD-BV, FTD-PNFA, FTD-SV) and for HCs of the MSCA cohort.

## Data Availability

All data produced in the present study are available upon reasonable request to the authors

https://adni.loni.usc.edu

https://memory.ucsf.edu/research-trials/research/allftd

https://www.ppmi-info.org/access-data-specimens/download-data

## Data availability

This study used publicly available data from three cohorts: the Alzheimer’s Disease Neuroimaging Initiative (ADNI), the Frontotemporal Lobar Degeneration Neuroimaging Initiative (NIFD), and the Parkinson’s Progression Markers Initiative (PPMI).

ADNI data are available from the Laboratory of Neuro Imaging (LONI) Image and Data Archive (IDA) at https://ida.loni.usc.edu, following registration and approval of a data use agreement.

NIFD data were obtained through the Frontotemporal Lobar Degeneration Neuroimaging Initiative, now continued as the ARTFL-LEFFTDS Longitudinal Frontotemporal Lobar Degeneration Study (ALLFTD). Data are available from the LONI IDA at https://ida.loni.usc.edu, following registration and approval of a data use agreement. Further information on the study is available at https://memory.ucsf.edu/research-trials/research/allftd.

PPMI data are available from the PPMI database at https://www.ppmi-info.org/access-data-specimens/download-data, following registration and approval of a data use agreement.

Anonymized data from the Amsterdam MS Cohort supporting the findings of this study are available from the corresponding author upon reasonable request, for research purposes only, subject to institutional and ethical approval.

## Acknowledgements

Data collection and sharing for the Alzheimer’s Disease Neuroimaging Initiative (ADNI) is funded by the National Institute on Aging (National Institutes of Health Grant U19 AG024904). The grantee organization is the Northern California Institute for Research and Education. In the past, ADNI has also received funding from the National Institute of Biomedical Imaging and Bioengineering, the Canadian Institutes of Health Research, and private sector contributions through the Foundation for the National Institutes of Health (FNIH) including generous contributions from the following: AbbVie, Alzheimer’s Association; Alzheimer’s Drug Discovery Foundation; Araclon Biotech; BioClinica, Inc.; Biogen; Bristol-Myers Squibb Company; CereSpir, Inc.; Cogstate; Eisai Inc.; Elan Pharmaceuticals, Inc.; Eli Lilly and Company; EuroImmun; F. Hoffmann-La Roche Ltd and its affiliated company Genentech, Inc.; Fujirebio; GE Healthcare; IXICO Ltd.; Janssen Alzheimer Immunotherapy Research & Development, LLC.; Johnson & Johnson Pharmaceutical Research &Development LLC.; Lumosity; Lundbeck; Merck & Co., Inc.; Meso Scale Diagnostics, LLC.; NeuroRx Research; Neurotrack Technologies; Novartis Pharmaceuticals Corporation; Pfizer Inc.; Piramal Imaging; Servier; Takeda Pharmaceutical Company; and Transition Therapeutics.

## SUPPLEMENTARY MATERIALS

**Supplementary Figure 1.**
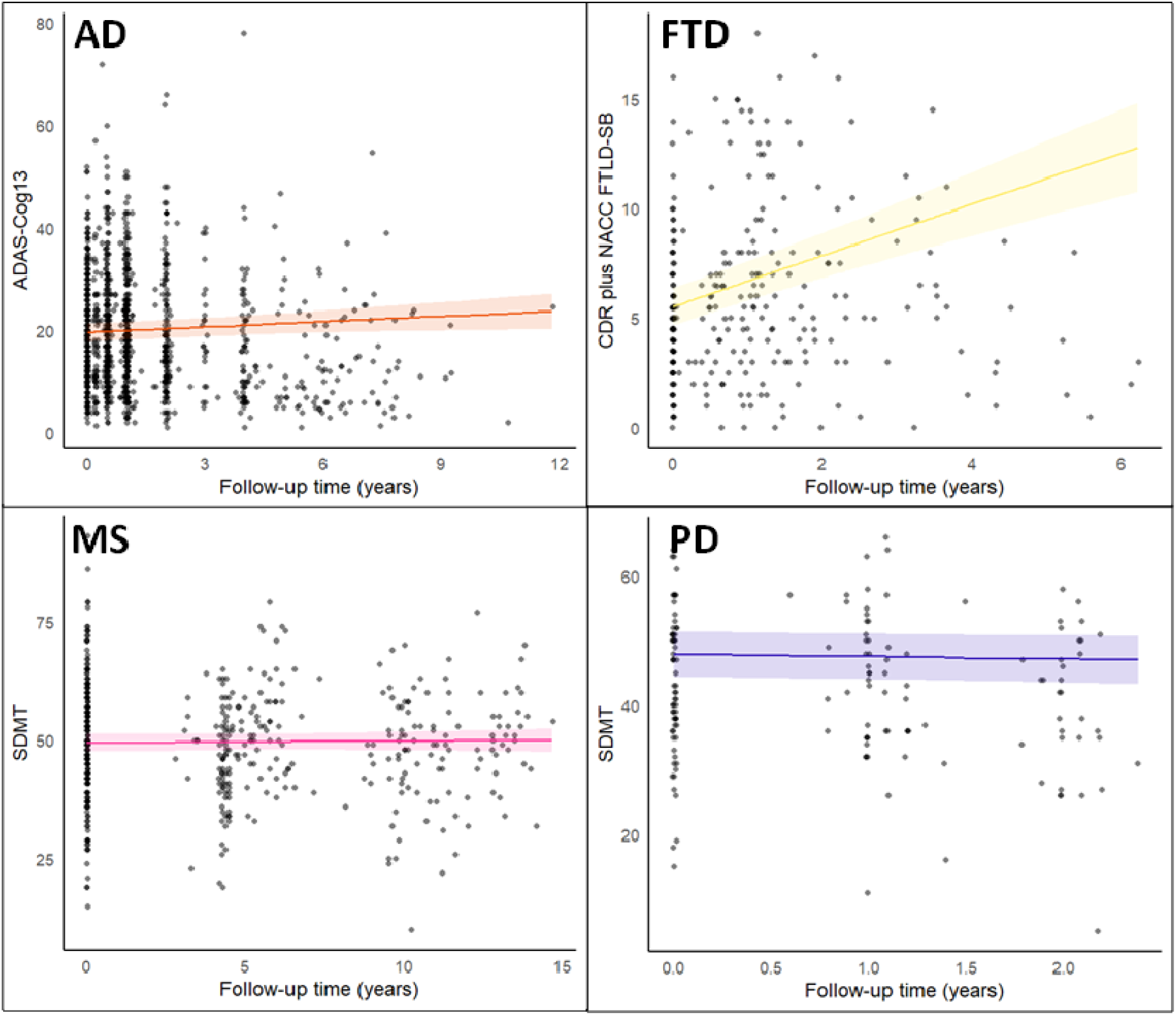
Cognitive change over time across diagnostic groups. Scatterplots showing the marginal effect of follow-up time on disease-specific cognitive measures in the AD (A), FTD (B), MS (C), and PD (D) patient groups. Linear fit lines are shown as solid lines (with corresponding 95% confidence interval shaded bands). All models were adjusted for age and sex.

**Supplementary Figure 2.**
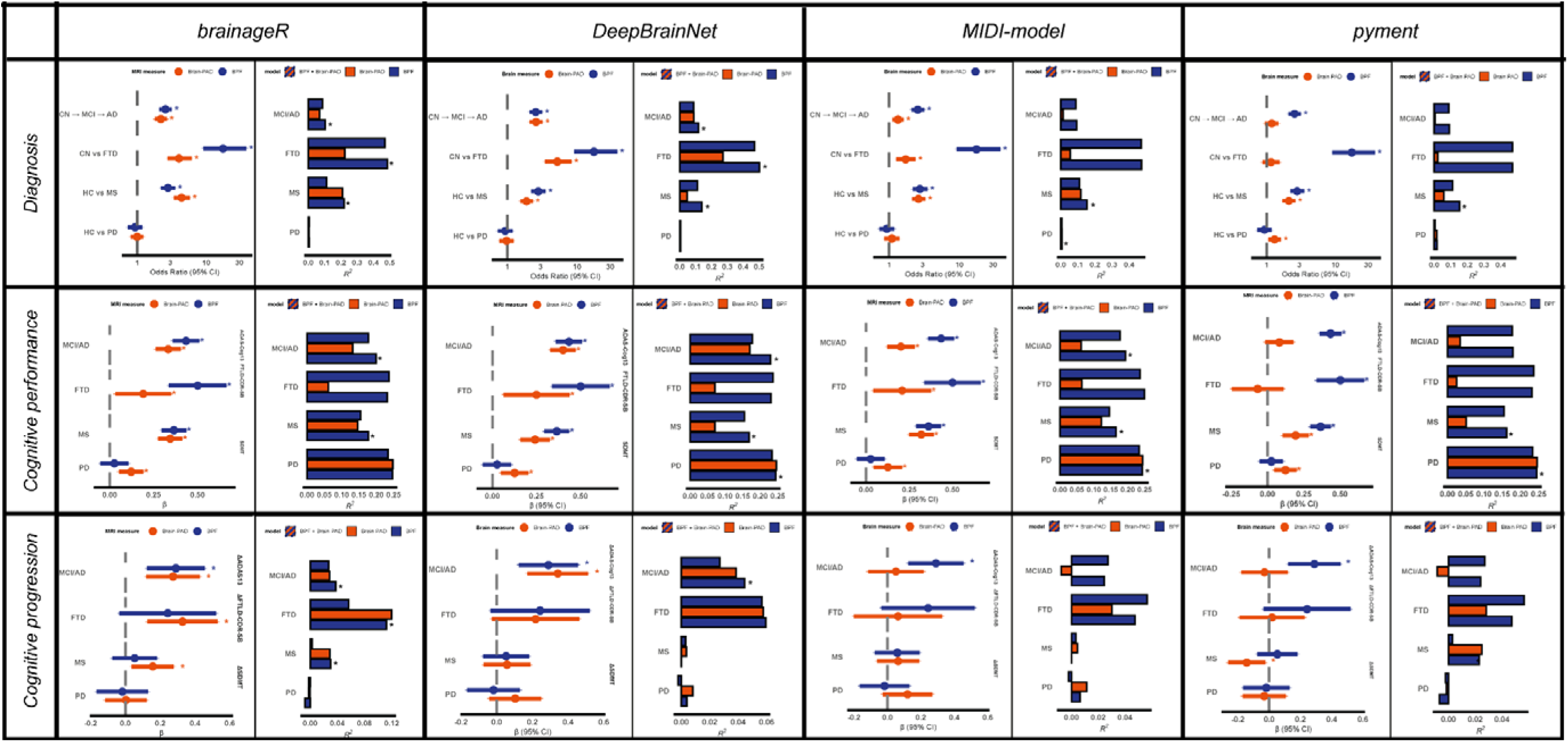
Clinical associations of BPF and brain-PAD values obtained from different brain age models. Figure showing the associations of BPF and brain-PAD computed with different models with diagnosis and cognitive performance and progression. Odds ratios (ORs) indicate the increased chances of belonging to the (more severe) patient group associated with one standard deviation change in the MRI measure. Standardized beta (β) values indicate changes in the dependent variable (in standard deviations) per one standard deviation change in the independent variable. To ease interpretation, signs of BPF and SDMT were flipped so that higher ORs or βs consistently indicate a stronger positive association between MRI-based neurodegeneration (i.e., higher brain-PAD or lower BPF) and the odds of belonging to the (more severe) patient group or cognitive severity/progression. All models were adjusted for age and sex. * indicates statistical significance.

**Supplementary Figure 3.**
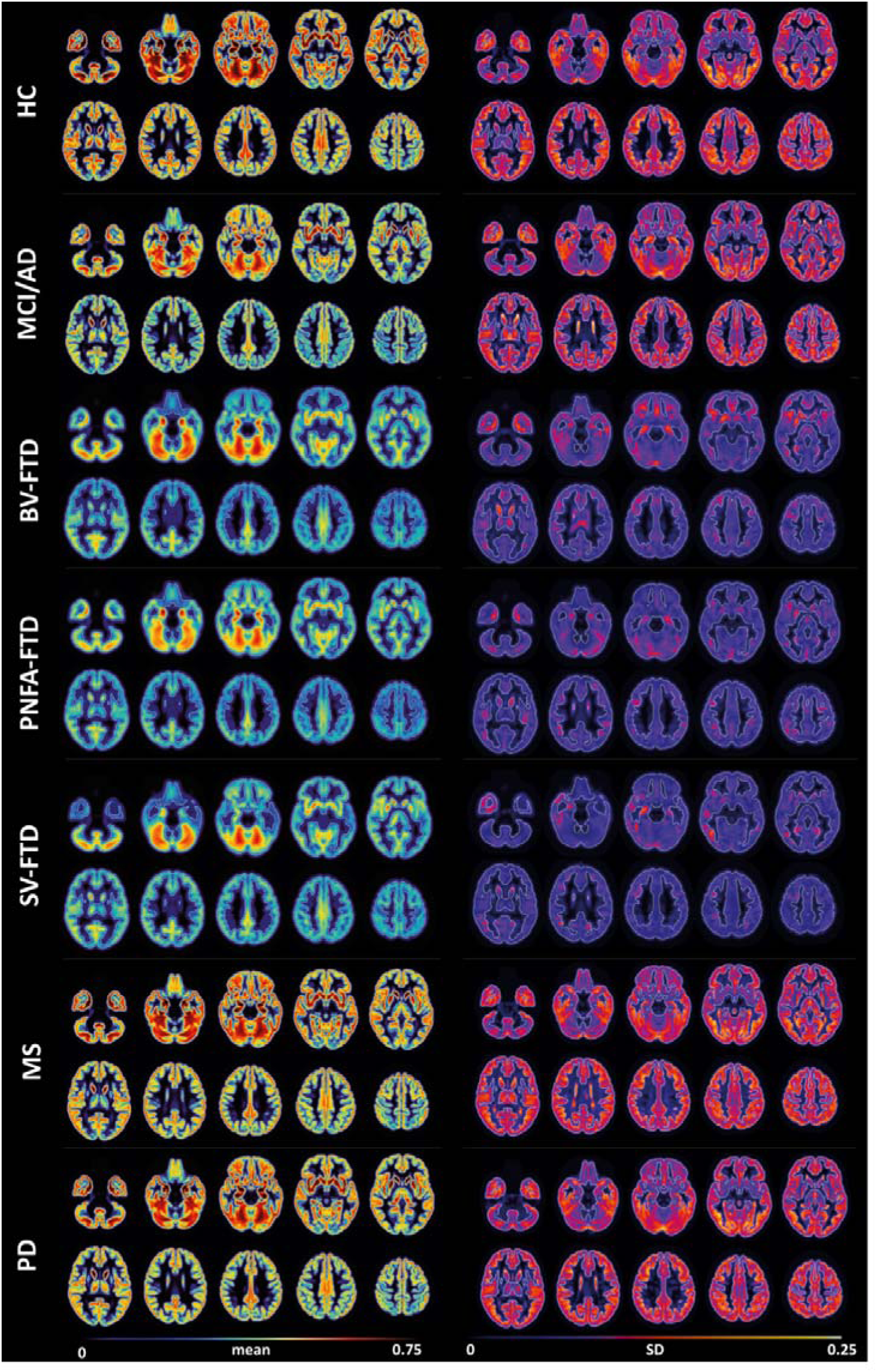
Voxel-level mean and SD values of preprocessed grey matter maps across diagnostic groups.

**Supplementary Figure 4.**
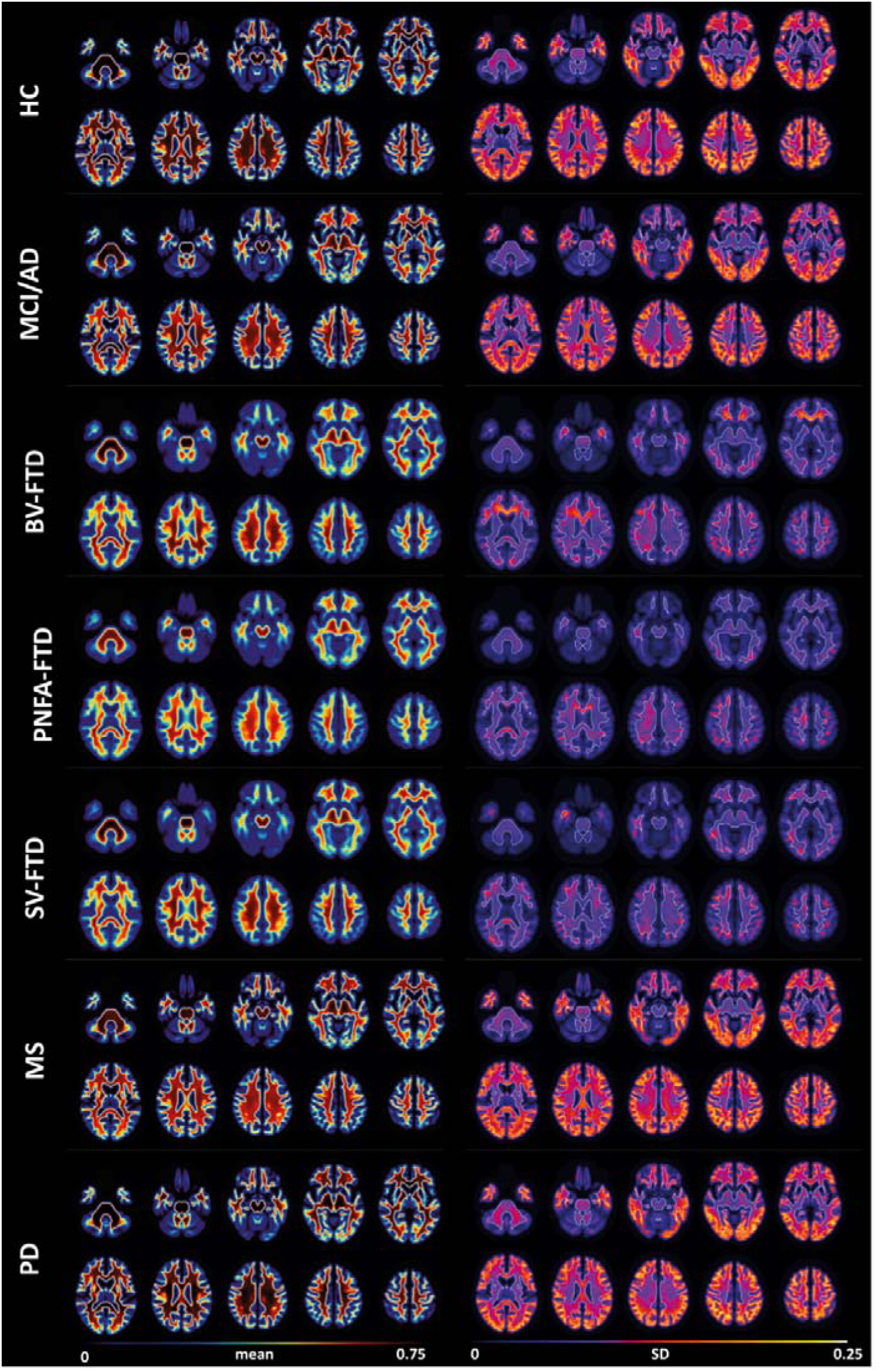
Voxel-level mean and SD values of preprocessed white matter maps across diagnostic groups.

**Supplementary Figure 5.**
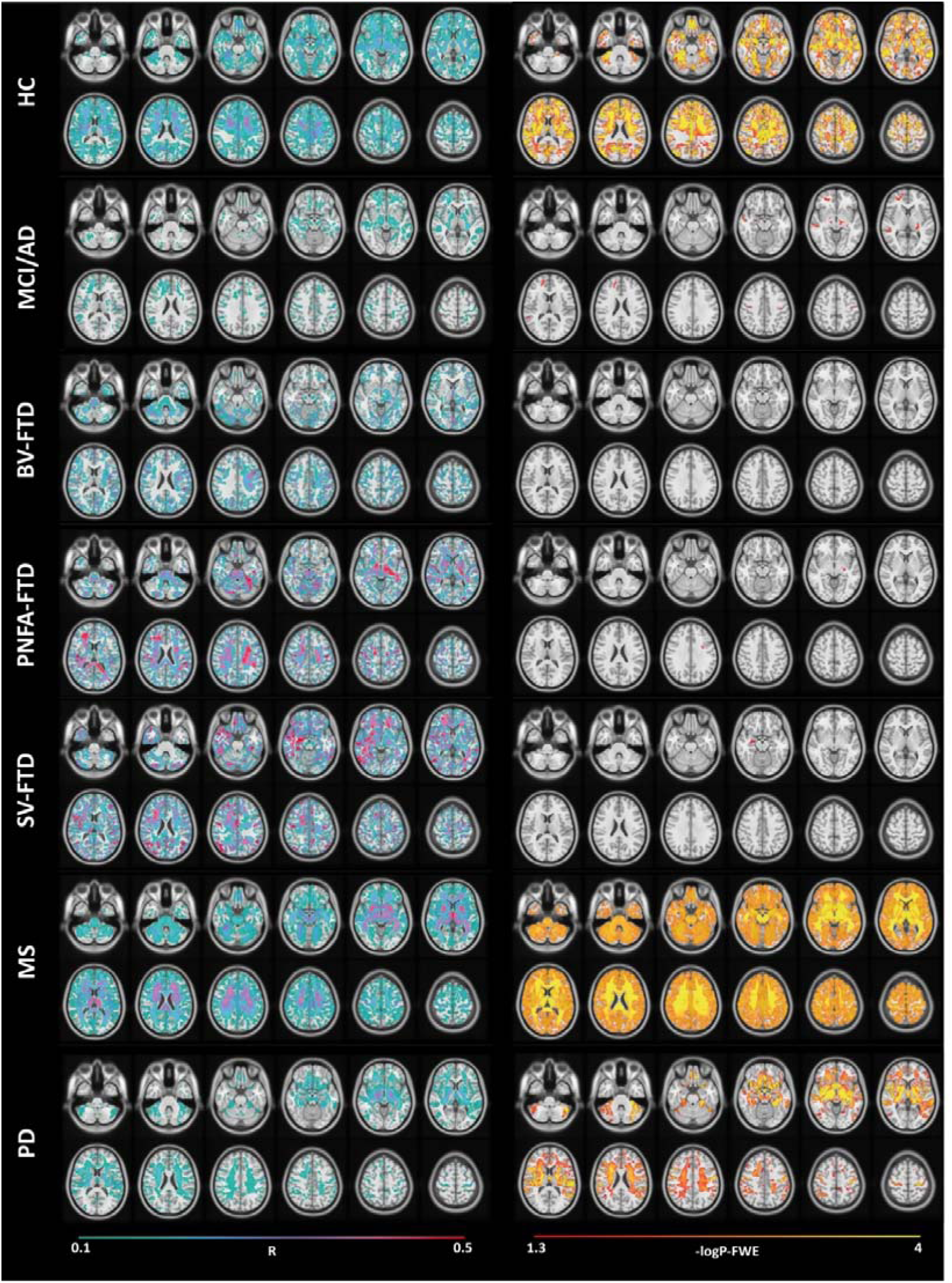
Voxel-wise associations between brain-PAD and brain tissue volumes, adjusted for BPF. Effect size (R, *sea green* to *red*) and thresholded statistical (−logP-FWE, *red* to *yellow*) maps of the negative relationship between brain-PAD and voxel-wise tissue volume are shown across diagnostic groups, superimposed on axial sections of a 3D T1-weighted template in standard space. All analyses were adjusted for age, sex, total intracranial volume, and brain parenchymal fraction (BPF).

